# Validated models for pre-test probability of stable coronary artery disease: a systematic review suggesting how to improve validation procedures

**DOI:** 10.1101/2020.11.27.20239301

**Authors:** Pierpaolo Mincarone, Antonella Bodini, Maria Rosaria Tumolo, Federico Vozzi, Silvia Rocchiccioli, Gualtiero Pelosi, Chiara Caselli, Saverio Sabina, Carlo Giacomo Leo

**Affiliations:** National Research Council, Institute for Research on Population and Social Policies, Brindisi (ITALY); National Research Council, Institute for Applied Mathematics and Information Technologies “Enrico Magenes”, Milano (ITALY); National Research Council, Institute of Clinical Physiology, Pisa (ITALY); National Research Council, Institute of Clinical Physiology, Lecce (ITALY)

**Author notes:** Corresponding author., - Complete address for correspondence National Research Council, Institute of Clinical Physiology, c/o Campus Ecotekne, Via Monteroni 73100 Lecce (ITALY). The first two authors equally contributed to the study.

**Keywords:** Coronary Artery Disease, Pre-test probability models, Validated models, Risk Assessment, Discrimination

## Abstract

An overuse of invasive and non-invasive anatomical testing for the diagnosis of coronary artery disease (CAD) affects patients’ and healthcare professionals’ safety, and the sustainability of Healthcare Systems. Pre-test probability (PTP) models can be routinely used as gatekeeper for initial patient management. Although with different positions, international organizations clearly underline the need for more information on the various risk factors acting as modifier of the PTP.

This systematic review addresses validation of PTP models adopting variables available at the first-line assessment of a suspected stable CAD. A comprehensive search has been done in MEDLINE®, HealthSTAR, and Global Health databases.

Nearly all the models considered in the 27 analysed papers include age, sex, and chest pain symptoms. Other common risk factors are smoking, hypertension, diabetes mellitus and dyslipidaemia. Only one model considers genetic profile. Reported AUCs range from 0.51 to 0.81. Relevant heterogeneity sources have been highlighted, such as the sample size, the presence of a PTP cut-off and the adoption of different definitions of CAD which can prevent comparisons of results. Very few papers address a complete validation, making then impossible to understand the reasons why the model does not show a good discrimination capability on a different data set.

We consequently recommend a more clear statement of endpoints, their consistent measurement both in the derivation and validation phases, more comprehensive validation analyses and the enhancement of threshold validations of PTP to assess the effects of PTP on clinical management.

## Introduction

Cardiovascular Diseases (CVDs) are the leading cause of mortality and morbidity worldwide with 422.7 million prevalent cases and 17.92 million deaths (one-third of all deaths) estimated in the most recent analysis of global burden of CVDs.^1^ Coronary artery disease (CAD) accounts for a large proportion of prevalent cases of CVDs after 40 years of age. CAD is one of the important causes of cardiovascular morbidity and mortality with a global estimation of 110.55 million prevalent cases and 8.92 deaths, which makes CAD the leading cause of death in the world.^1^

Stable CAD is most commonly caused by atherosclerotic coronary artery narrowing and is characterized by episodes of reversible myocardial demand/supply mismatch, related to ischaemia or hypoxia, which are usually inducible by exercise, emotion or other stress and commonly associated with transient chest discomfort (stable angina pectoris).^2,3^ Stable CAD diagnosis is established through non-invasive functional and/or anatomical testing,^2,3^ and invasive coronary angiography (ICA).^2^ Preventive medication plus symptomatic medical management and/or revascularization are the current treatment strategies for established stable CAD.^2,3^

To limit the risk of inappropriate examinations, with its consequences on patients’ and healthcare professionals’ safety, and economic sustainability of Healthcare Systems,^4–7^ eligibility to diagnostic testing is established through models that predict a pre-test probability (PTP) of coronary artery disease (CAD). Since the introduction of the Diamond-Forrester model (DFM)^8^ and the Duke Clinical Score (DCS)^9^ several alternative PTP models have been proposed and recommended in guidelines for stable symptomatic subjects.^3,10^ Recent updates in the European scenario stressed the overestimation flaw of such models. As a consequence, the UK National Institute for Health and Care Excellence (NICE) has preferred a simpler identification of anginal chest pain or abnormal resting electrocardiogram (ECG) as a gatekeeper to Coronary computed tomography angiography (CCTA).^11^ However, the performance of CCTA for the diagnosis of obstructive CAD is not significantly influenced by chest pain symptoms and angina is more than a mere biological phenomenon which requires specific attentions especially in women.^12,13^ The European Society of Cardiology (ESC) updated guideline determines PTP from the stratified prevalence of CAD in a contemporary cohort, instead of recurring to a prediction model as in the past. These new estimated risks are noticeably lower compared to the previous ones possibly suggesting underestimation when applied to different populations, as also recently stated by Bing and colleagues.^14^ US Experts are debating on whether adopting the NICE diagnostic approach or keeping on using PTP.^15,16^ To face the flaws on available PTP models highlighted by NICE and ESC, these organizations clearly underline the need for more information on the various risk factors acting as modifier of the PTP, especially in the low probability range,^10^ and for the development and validation of new scores addressing outstanding uncertainties in the estimation of the PTP of CAD.^11^

This review provides several new contributions to the actual debate on how to ameliorate the PTP models as it focuses on external validation mainly^17^ identifies the best results and characterizes the best procedures in terms of significant predictive variables, discriminatory ability and methods completeness. Moreover, the review highlights some key issues that could be further improved in the development and validation phases, to increase decision making capability.

## 2. The systematic review: how it works

This systematic review conforms to the PRISMA statement;^18^ the protocol was registered in PROSPERO (CRD42019139388).^19^

### 2.1 Study inclusion and exclusion criteria

Inclusion and exclusion criteria were developed to identify studies that validated PTP models of stable obstructive CAD (as a binary outcome) anatomically determined through either ICA or CCTA. Reasons of exclusion were: (i) acute coronary syndrome, unstable chest pain, a history of myocardial infarction or previous revascularisation; (ii) models that included a diagnostic procedure that do not reflect the usual practices of the first-line assessment;^3,10^ (iii) models based on a single predictive variable; (iv) lack of clearly stated discrimination power. Unlike previous works,^20^ external validation was primarily considered. We also included internal validation but limited it to *k-*fold cross-validation as a technique inspired by the same purposes of external validation. Moreover, papers referring to Machine learning-based PTP models have been excluded as considered in a recent review focusing on CAD diagnosis by ML with aims close to ours.^21^ Only full papers were retained because other publications, e.g., letters to editors, conference proceedings, et cetera, are usually not assessed for study quality. Only articles published in English and Italian were considered.

### 2.2 Searches

The databases Global Health, Healthstar and MEDLINE® were systematically searched (CGL, PM) on 22 April 2020 using several keywords, including: angina pectoris, chest pain, coronary artery disease, coronary heart disease, coronary stenosis, stratification score, likelihood function, predictive model, pre-test probability, coronary angiography, cardiac catheterisation and computed tomography angiography. Full electronic search strategy is reported in Additional file 1. Citation searches were also performed on reference lists of definitively included studies.

### 2.3 Study selection

A multidisciplinary working team was composed. Eligibility screening was performed independently in an unblinded standardized manner by all the reviewers. Preliminary screening was performed using Abstrackr^22^ based on title and abstract with each paper assessed by two randomly assigned reviewers. Selected papers were assessed based on full text. Disagreements were resolved by consensus.

### 2.4 Data extraction strategy

A data collection form was developed by three authors (A.B, CGL, PM) and filled by reviewers independently. Three authors (A.B, CGL, PM) reviewed the final form for internal consistency. Each selected paper was assigned for data extraction to the statistician (AB) and two randomly selected reviewers.

### 2.5 Study quality assessment

The quality assessment of included studies conforms to QUADAS-2 and was performed by four reviewers (A.B, CGL, PM, MRT).^23^ Due to the previously described features (i)-(iv), we considered that the eligible works did not raise applicability concerns.

### 2.6 Data synthesis and presentation

The performances of prediction models can be summarised using several methods and indices, and the area under receiver operating characteristic curve (AUC) is certainly the best-known. Sensitivity and specificity also describe the discrimination capability of the model for a given cut-off and thus provide an indication of clinical usefulness.

For the purposes of generalisation of a PTP model to populations that differ from the development population study, the computation of performance indexes is not sufficient because a lower performance is usually expected.^17,24^ Therefore, we also noted whether more extended validation procedures were performed in order to properly apply a model to new populations.

## 3. Validated PTP models: main results

### 3.1 Study selection

A total of 5,711 studies were identified (3 through reference lists of included studies) and 2,685 different abstracts were screened. Out of the 71 relevant full-texts assessed for eligibility, 27 were finally included (Figure 1).

**Figure 1:**
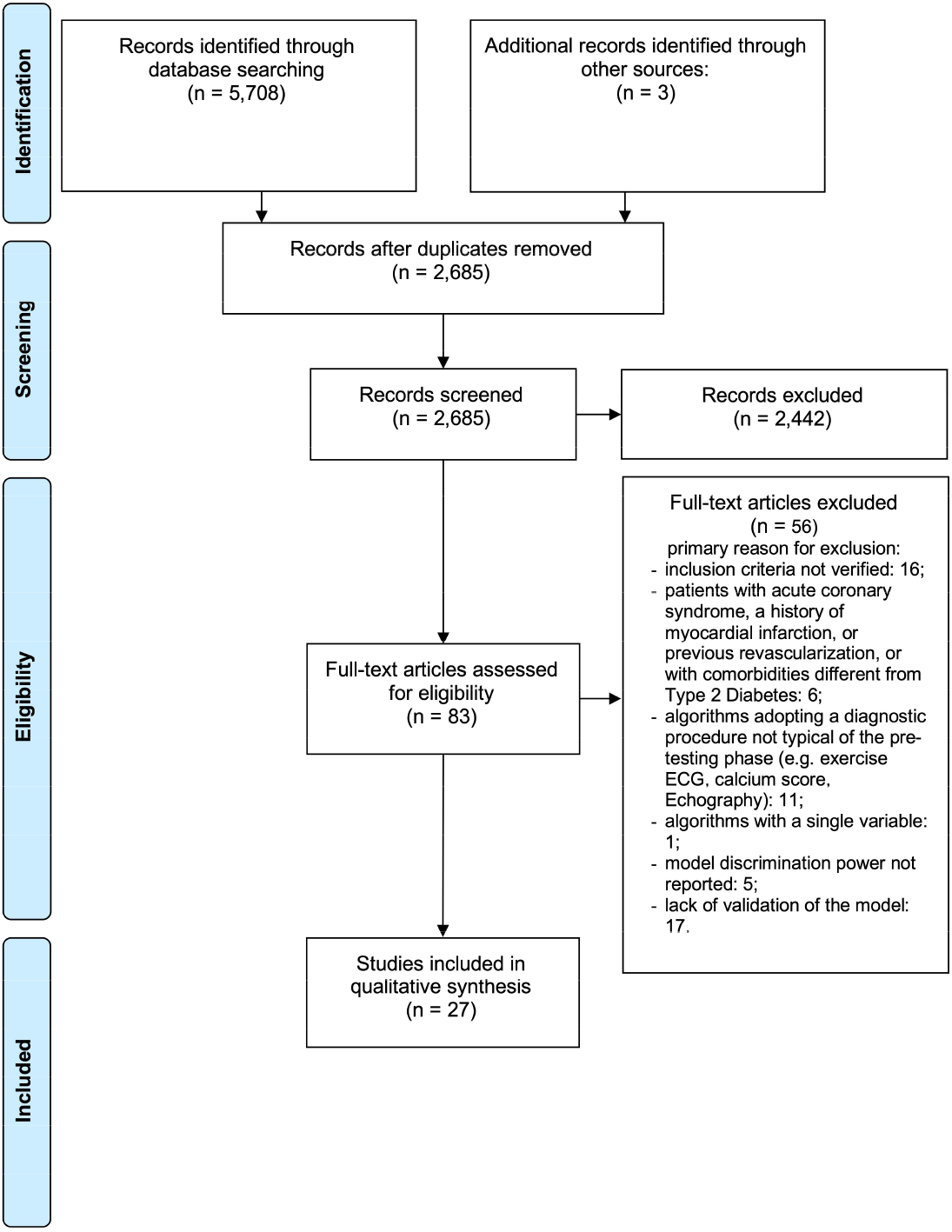
Search and selection process for systematic review according to PRISMA.

### 3.2 Study characteristics

Table 1 summarises the selected studies in terms of model name, geographical location, population recruitment criteria. Sometimes the same model is referenced with different names across the papers, then Table 1 indicates the original name and the one we adopted here.

**Table 1:** Characteristics of the studies on PTP for CAD.

| Study | Models / scores | Study Centers | Population | Exclusion criteria |
| --- | --- | --- | --- | --- |
|  |  |  | Inclusion criteria |  |
| Adamson PD, 2018a<sup>25</sup> | DFM/CASS<br>uDFM | 1) Multicenter PROMISE trial, US & Canada | See PROMISE. Randomised to receive CCTA as non-initial non-invasive test. | See PROMISE & SCOT-HEART. Known CAD |
|  |  | 2) Multicenter SCOT-HEART trial, Scotland (UK) | See SCOT-HEART. Randomized to the CCTA intervention arm. |  |
| Adamson PD, 2018b<sup>26</sup> | uDFM (Baseline CADC model, in text)<br>uDFM-cTn (Baseline CADC model with the addition of troponin, in text) | Odense University Hospital, Denmark | Clinical stable prospectively enrolled patients with suspected angina pectoris scheduled for either ICA or CCTA<sup>27</sup> | Suspected acute coronary syndrome. To avoid potential confounding effects on the biomarkers measured, patients with established atherosclerotic manifestations, including an abnormal 12-lead rest electrocardiogram, were excluded: known ischemic heart disease, prior ischemic stroke or transitory ischemic attack, known peripheral artery disease (n = 10), and p-creatinine >200 mmol/L. CCTA not performed or of poor technical quality, lack of informed consent, missing hs-cTnI measure or personal history.<sup>27</sup> |
| Almeida J, 2016<sup>28</sup> | CADC-Clin (CAD Consortium 2, in text)<br>DCS<br>uDFM (CAD Consortium 1, in text) | Single center in Southwestern Europe | Patients with chest pain and suspected CAD referred to ICA | Patients with a history of CAD, acute coronary syndrome, or coronary revascularization |
| Baskaran L, 2018<sup>29</sup> | CADC-Clin<br>CONFIRM score | Multicenter SCOT-HEART | See SCOT-HEART. Randomized to the CCTA intervention arm and with information on all | See SCOT-HEART. Known CAD |

| Study | Models / scores | Study Centers | Population |  |
| --- | --- | --- | --- | --- |
|  |  |  | Inclusion criteria | Exclusion criteria |
|  | uDFM | trial, Scotland (UK) | variables needed for the analysis. |  |
| Bittencourt MS, 2016<sup>30</sup> | CADC-Basic<br>CADC-Clin<br>uDFM (Diamond and Forrester score, in text) | Massachusetts General Hospital; Brigham and Women's Hospital (Massachusetts, USA) | Subjects ≥ 18 years who underwent CCTA for suspect of CAD | Patients who were missing any of the clinical information needed to calculate the pretest probability, who had nondiagnostic CCTA images, who had incomplete follow-up information; with congenital heart disease, heart transplantation, or prior CAD, defined as prior percutaneous coronary interventions, coronary artery bypass graft surgery, or MI |
| Daniels SE, 2014<sup>31</sup> | Corus® CAD (Gene Expression score – GES, in text) | Multicenter PREDICT trial US | See PREDICT | See PREDICT. Diabetic patients |
| Edlinger M, 2017<sup>32</sup> | CADC-Clin | University Clinic of Cardiology at Innsbruck (Austria) | Patients were 18 years of age or older with chest pain or symptoms suggestive of CAD (predominantly dyspnoea) and/or non-invasive evidence of CAD referred for elective ICA. | 1) an elective ICA before or after heart transplantation, 2) an elective ICA prior to solid organ transplantation, 3) an elective ICA before heart valve repair or replacement, or with valvular heart disease as leading clinical diagnosis, 4) an isolated right heart catheterisation, 5) an electrophysiological procedure (pace-maker implantation or catheter ablation) as leading clinical indication, 6) an elective ICA because of a known or suspected congenital heart disease as leading clinical diagnosis (e.g., atrial septal defect, ventricular septal defect or patent foramen ovale), or 7) when referred for other reasons (like myocardial biopsy, aortic aneurysms, |
|  |  |  | Inclusion criteria | Exclusion criteria |
|  |  |  |  | myxoma, endocarditis or prior failed angiography). History of myocardial infarction. |
| Ferreira AM, 2016<sup>33</sup> | uDFM (Modified DF, in text)<br>CADC-Clin (CAD consortium 2, in text)<br>CONFIRM score | Unspecified, Portugal | Patients undergoing CCTA for the evaluation of CAD | Age <30 years; known CAD; suspected acute coronary syndrome; preoperative assessment; known left ventricular systolic dysfunction; asymptomatic patients (typically referred after a positive screening exercise test); symptoms other than chest pain. Patients with suspected CAD who were scheduled to undergo CCTA but had the procedure halted due to a high coronary artery calcium (CAC) Agatston score. A threshold of 400 was used as a general guideline for withholding CCTA in these circumstances, but the decision was ultimately left to the performing physician, taking into consideration the clinical context and the distribution of calcium in the coronary tree. |
| Fordyce CB, 2017<sup>34</sup> | PROMISE minimal risk model<br>(The originally published version has been subsequently corrected online, see Fordyce CB, 2018<sup>35</sup>) | Multicenter PROMISE trial, US & Canada | See PROMISE. Patients assigned to anatomic testing | See PROMISE |
| Fujimoto S, 2014<sup>36</sup> | DCS<br>K-score | Multicenter, Japan | Suspected CAD | Patients with known CAD, showing poor image quality and patients with un-assessable segments due to severe calcification |
| Genders | DFM | 14 European | Patients aged 30-69 with stable chest pain | Patients meeting the following criteria: (i) acute coronary syndrome |
|  |  |  | Inclusion criteria | Exclusion criteria |
| TSS,<br>2011<sup>37</sup> |  | centers | (typical, atypical, or non-specific chest pain) and if ICA performed. | or unstable chest pain, (ii) history of myocardial infarction or previous revascularization (percutaneous coronary intervention or coronary artery bypass graft surgery), and (iii) no informed consent. |
|  | uDFM | Erasmus Medical Center, Rotterdam, the Netherlands<sup>38</sup> | Patients with stable chest pain and no history of CAD<sup>38</sup> | Not undergoing CCTA or ICA |
| Genders<br>TSS,<br>2012<sup>39</sup> | DCS | Multicenter EU and US | Stable chest pain, referred for catheter based or CT based coronary angiography | Acute coronary syndrome, unstable chest pain, history of myocardial infarction or previous revascularization or no informed consent. |
|  | CADC-Basic<br>CADC-Clin | Multicenter EU and US | Stable chest pain, referred for catheter based or CT based coronary angiography | Acute coronary syndrome, unstable chest pain, history of myocardial infarction or previous revascularization or no informed consent. |
| Genders<br>TSS,<br>2018<sup>40</sup> | CADC-Basic<br>CADC-Clin | Multicenter PROMISE trial, US & Canada | See PROMISE Trial for the main criteria.<br>Patients assigned to anatomic testing | See PROMISE Trial for the main criteria |
| Jensen JM,<br>2012<sup>41</sup> | CORSCORE<br>DCS<br>DFM<br>Morise score<br>uDFM | Lillebælt Hospital Vejle, Denmark | Patients with chest pain indicative of CAD referred for ICA | Unstable angina or previous coronary intervention |
| Min JK,<br>2015<sup>42</sup> | CONFIRM score (Integer-based risk model, in text) | United States, Canada, South | Patients $\geq 18$ years old referred to CCTA for suspected stable CAD (CONFIRM trial<sup>43</sup>) | Patients with prior coronary revascularization or MI, asymptomatic, missing data |
|  |  |  | Inclusion criteria | Exclusion criteria |
|  |  | Korea and Austria (4 out of 5 sites of the Phase II of CONFIRM trial<sup>43</sup>) |  |  |
| Pickett CA, 2013<sup>44</sup> | DFM/CASS<br>Morise score | Walter Reed Army Medical Center, Washington USA | Patients referred for CCTA | Known CAD |
| Rademaker AA, 2014<sup>45</sup> | DCS<br>DFM<br>Morise score (New score, in text)<br>uDFM | VU University Medical Center, Amsterdam, The Netherlands | Symptomatic women undergoing evaluation for CAD and referred for CCTA | Prior history of CAD (percutaneous coronary intervention, coronary artery bypass graft surgery, or previous myocardial infarction), or absolute or relative contraindications for CCTA such as (i) significant severe arrhythmia; (ii) pregnancy; (iii) renal insufficiency (glomerular filtration rate<45 ml/min); (iv) known allergy to iodinated contrast material. |
| Rosenberg S, 2010<sup>46</sup> | Corus® CAD (Gene expression test, in text)<br>Expanded clinical model score<br>DFM/CASS | Multicenter<br>PREDICT trial<br>US | See PREDICT | See PREDICT. Diabetes |
| Teresa G, | CADC-Basic | 1 center in US | >18 years old evaluated in the Emergency | Known CAD, defined as history of acute myocardial infarction, |
|  |  |  | Inclusion criteria | Exclusion criteria |
| 2018<sup>47</sup> | CADC-Clin | | Department of a major academic tertiary university hospital for chest pain, using CCTA as a primary diagnostic modality | percutaneous intervention, coronary artery bypass graft, or evidence of CAD by either anatomical (CCTA or cardiac catheterization) or functional tests (positive stress test). Hemodynamically or clinically unstable patients, patients with ST segment changes or positive cardiac troponin ($>0.04\text{ng/ml}$), impaired renal function ($\text{eGFR} < 50\text{ml/min/1.73m}^2$), tachycardia, or contraindication to nitroglycerin or iodinated contrast. Inadequate documentation on Chest pain characteristics, repeat CCTAs, unavailable calcium score and non-diagnostic exam. |
| Thomas GS 2013<sup>48</sup> | Corus® CAD (GES, in text)<br>DFM<br>Morise score | Multicenter COMPASS trial, US | See COMPASS | See COMPASS |
| Voora D, 2017<sup>49</sup> | Corus® CAD | Multicenter PROMISE trial, US & Canada | See PROMISE. Patients assigned to anatomic testing | See PROMISE. Diabetes. RNA sample not passing quality control. |
| Voros S, 2014<sup>50</sup> | Corus® CAD (GES, in text)<br>DFM | Multicenter PREDICT US and COMPASS US trials | See PREDICT and COMPASS. | See PREDICT and COMPASS. Diabetes excluded from PREDICT cohort. |
| Wang M 2018<sup>51</sup> | CONFIRM score | Not specified, China | Patients who underwent CCTA for stable chest pain and with 0 or 1 risk factors among smoking, hypertension, diabetes and | Acute coronary syndrome, previous CAD or coronary revascularization, un-assessable segments due to motion artifact, atrial fibrillation, aortic disease, New York Heart Association class |
|  |  |  | Inclusion criteria | Exclusion criteria |
|  |  |  | hyperlipidemia | III or IV heart failure, age > 90 years old, pacemaker leads or missing data |
| Winther S, 2019<sup>52</sup> | uDFM<br>CADC-Basic<br>CADC-Clin | Multi-center<br>Dan-NICAD<br>trial, Denmark | Patients without known CAD referred to CCTA due to a history of symptoms suggestive of CAD | Age <40; previous coronary revascularization or MI; unstable angina pectoris; estimated glomerular filtration rate <40mL/min; pregnancy; and contraindication for iodine-containing contrast medium, magnetic resonance imaging, or adenosine (severe asthma, advanced atrioventricular block, or critical aortic stenosis). |
| Yang Y, 2015<sup>53</sup> | High Risk Anatomy (HRA) score | Multicenter<br>CONFIRM<br>trial,<sup>43</sup> North<br>America,<br>Europe and<br>Asia<br>University of<br>Ottawa Heart<br>Institute<br>Cardiac CT<br>registry | Patients ≥18 years old referred to CCTA for suspected stable CAD (CONFIRM trial)<sup>43</sup> | Documented CAD, history of MI, coronary revascularization, cardiac transplantation, congenital heart disease |
|  | uDFM | Multicenter<br>CONFIRM<br>trial,<sup>43</sup> North<br>America,<br>Europe and | Patients ≥18 years old referred to CCTA for suspected stable CAD (CONFIRM trial)<sup>43</sup> | Documented CAD, history of MI, coronary revascularization, cardiac transplantation, congenital heart disease |
|  |  |  | Inclusion criteria | Exclusion criteria |
|  |  | Asia |  |  |
| Zhang Y, 2019<sup>54</sup> | DCS<br>uDFM | Tianjin Chest Hospital,<br>Tianjin, China | Patients with stable chest pain and referred for CCTA | Acute coronary syndrome, previous CAD or coronary revascularization (percutaneous coronary intervention or coronary artery bypass grafting), impaired renal function (serum creatinine > 120 µmol/l), New York Heart Association class III or IV heart failure, atrial fibrillation, aortic disease, age more than 90 years, or patients with un-assessable segments because of artefact |
| Zhou J, 2017<sup>55</sup> | CADC-Clin (Genders clinical model, in text)<br>DCS<br>uDFM | Not specified, China | Patients who underwent CCTA for stable chest pain | Acute coronary syndromes, previous CAD or coronary revascularization (percutaneous coronary intervention or coronary artery bypass grafting), patients with un-assessable segments due to motion artefact, atrial fibrillation, aortic disease, New York Heart Association class III or IV heart failure, age > 90 years, presence of pacemaker leads or missing data. |

The trials COMPASS, CONFIRM, PREDICT, PROMISE and SCOT-HEART were considered in several studies, and thus their main characteristics are fully reported in Additional File 2

Studies are mainly conducted in North America ^30,31,34,40,44,46–50^ or Europe^26,28,29,32,33,37,41,45,52^.

The Updated Diamond-Forrester model (uDFM),^25,26,53–55,28–30,33,37,41,45,52^ and the CAD consortium clinical model (CADC-Clin)^28–30,32,33,39,40,47,52,55^ are the most assessed models.

The quality of included studies is generally high due to the specific review question and adopted eligible criteria. Nevertheless, risk of bias arises from a few specific issues. A few validation studies^31,37,39,46,50^ do not declare that they enrolled only consecutive or random samples of patients. With respect to the index test, only one work adopted an optimal discriminating threshold in addition to pre-specified ones.^50^ Application of CCTA as a reference test yields a risk of bias in many studies^25,34,37,39,40,42,45,49,53^ that do not report measures against misclassification of the test results. Finally, in four works^26,39,40,48^ patients did not receive the same reference test for the diagnosis of stable CAD. A graphical summary of the risk of bias is reported in Additional File 3.

### 3.3 Predictive variables and discrimination capability

As shown in Table 2, the identified models can be classified into two broad classes: basic models, including the DFM (based on age, sex and chest pain) and its updates, and clinical models, including the DCS and the models that extend the DFM by adding a few, mainly traditional,^56^ risk factors. Within this quite classical framework, the Corus® CAD model is distinguished by relating CAD in nondiabetic patients to the expression levels of a set of genes.

**Table 2:** PTP models’ variables.

| Macro categories | Predicting variables | Model/Score |  |  |  |  |  |  |  |  |  |  |  |  |  |  |
| --- | --- | --- | --- | --- | --- | --- | --- | --- | --- | --- | --- | --- | --- | --- | --- | --- |
|  |  | CADC-basic | CADC-Clin | CONFIR M score | CORSCO RE | Corus® CAD | DCS | DFM | DFM/CA SS | Expanded clinical model score | K-score | HRA score | Morise score | PROMIS E Minimal Risk model | uDFM | uDFM-cTn |
|  |  | 30,39,40,47,52 | 28– | 29,33,42,51 | 41 | 31,46,48–50 | 28,36,39,41,45 | 37,41,42,45,48 | 25,44,46 | 46 | 36 | 53 | 41,44,45,48 | 34 | 25,26,53– | 26 |
|  |  |  | 30,32,33,39,40 |  |  |  | ,54,55 | ,50 |  |  |  |  |  |  | 55,28– |  |
|  |  |  | ,47,52,55 |  |  |  |  |  |  |  |  |  |  |  | 30,33,37,41,45 |  |
|  |  |  |  |  |  |  |  |  |  |  |  |  |  |  | ,52 |  |
| Demography | Age | √ | √ | √ | √ | √ | √ | √ | √ | √ | √ | √ | √ | √ | √ | √ |
|  | Sex | √ | √ | √ | √ | √ | √ | √ | √ | √ | √ | √ | √ | √ | √ | √ |
|  | Race |  |  |  |  |  |  |  |  | √ |  |  |  | √ |  |  |
| Medical history | Diabetes mellitus |  | √ | √ |  |  | √ |  |  |  | √ | √ | √ | √ |  |  |
|  | Hypertension |  | √ | √ | √ |  |  |  |  | √ | √ | √ | √ | √ |  |  |
|  | Previous MI |  |  |  | √ |  | √ |  |  |  |  |  |  |  |  |  |
|  | Cerebral Infarction |  |  |  |  |  |  |  |  |  | √ |  |  |  |  |  |
|  | Peripheral vascular disease |  |  |  |  |  |  |  |  |  |  | √ |  |  |  |  |
| Clinical presentation/physical | Chest pain | √ | √ | √ | √ |  | √ | √ | √ | √ | √ | √ | √ |  | √ | √ |
|  | Abnormal ECG |  |  |  |  |  | √ |  |  |  |  |  |  |  |  |  |
|  |  | CADC-basic | CADC-Clin | CONFIRM score | CORSCO RE | Corus® CAD | DCS | DFM | DFM/CA SS | Expanded clinical model score | K-score | HRA score | Morise score | PROMISE Minimal Risk model | uDFM | uDFM-cTn |
| examination | Obesity |  |  |  |  |  |  |  |  |  |  |  | √ |  |  |  |
|  | Smoking |  | √ | √ | √ |  | √ |  |  |  | √ | √ | √ | √ |  |  |
|  | Family history of CAD |  |  | √ |  |  |  |  |  |  |  | √ | √ | √ |  |  |
|  | Other (specify) |  |  |  | Medically treated hyperchol. |  |  |  |  | Medically treated hyperchol. |  |  |  | Symptoms related to physical or mental stress |  |  |
| Bio-Chemistry | HDL cholesterol |  |  |  |  |  |  |  |  |  |  |  |  | √ |  |  |
|  | Dyslipidaemia |  | √ |  |  |  | √ |  |  | √ | √ | √ | √ | √ |  |  |
|  | Oestrogen status |  |  |  |  |  |  |  |  |  |  |  | √ |  |  |  |
|  | Gene expression |  |  |  |  | √ |  |  |  |  |  |  |  |  |  |  |
|  | Troponin |  |  |  |  |  |  |  |  |  |  |  |  |  |  | √ |
| Others |  |  |  |  |  |  |  |  |  | Aspirin, anti- |  |  |  |  |  |  |
|  |  | CADC-basic | CADC-Clin | CONFIRM score | CORSCO RE | Corus® CAD | DCS | DFM | DFM/CA SS | Expanded clinical model score | K-score | HRA score | Morise score | PROMISE Minimal Risk model | uDFM | uDFM-cTn |
|  |  |  |  |  |  |  |  |  |  | platelet, ACE inhibitor use, systolic blood pressure |  |  |  |  |  |  |
| Derivation method |  | Log | Log | Cox proportional hazards models | Log | Score derived by a Ridge regression | Log | Conditional probability analysis * | Log | Log | Log | Score derived by a multivariable Log | Score derived by a Log | Log | Log | Log |

\* In Genders, 2011,<sup>37</sup> to unravel the implicit coefficients of the predictors in this model, the authors performed a weighted linear regression on the log odds of the DF predictions per subgroup

All the models were derived by logistic regression. Exceptions are: DFM, derived by a conditional probability analysis in the late 1970s; Corus® CAD, obtained through Ridge regression; CONFIRM score, developed to predict adverse clinical events by fitting a Cox proportional hazards model and subsequently validated for diagnosis of CAD. Cross-validation^39^ and split-sample^34,46^ have been used in a few cases only.

Predictors were classified into four macro-areas: demography, medical history, clinical presentation/physical examination and biochemistry. The demographic macro-area is present in all models with the variables age and sex, while race is only included in the Expanded clinical model and PROMISE Minimal Risk model. The most used variables in the medical history macro-area are diabetes mellitus and hypertension. The clinical presentation/physical examination macro-area is present in all but the Corus® CAD models.

Only the Corus® CAD and PROMISE Minimal Risk models do not include chest pain. The most used variable in the biochemistry macro-area is dyslipidaemia. The other risk factors are model-specific: gene expression (Corus® CAD), oestrogen status (Morise score), high-density lipoprotein cholesterol (PROMISE Minimal Risk model) and the high-sensitivity cardiac troponin (uDFM-cTn).

Finally, Table 3 reports the overall picture of the PTP discriminatory abilities in the validation studies.

**Table 3:**
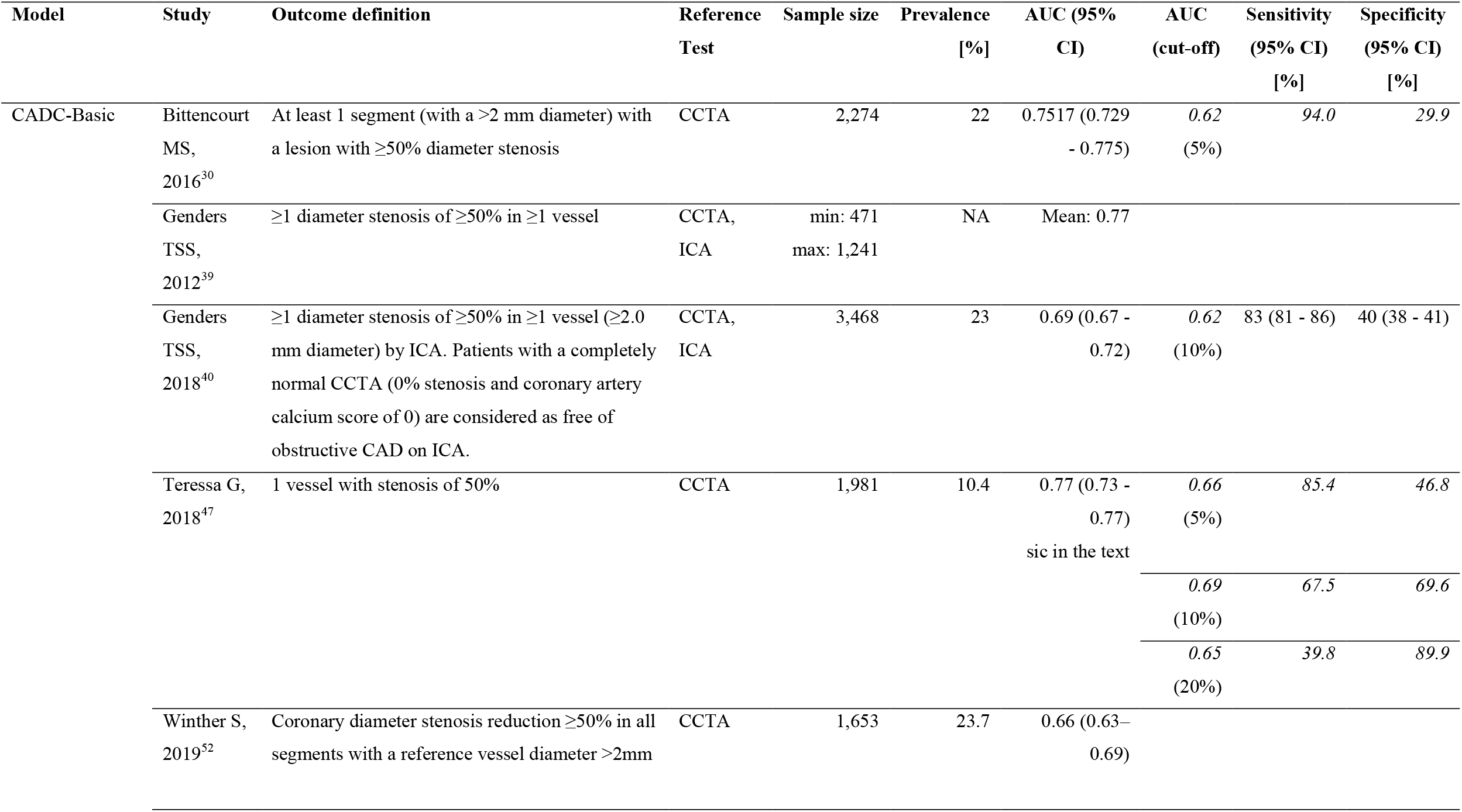
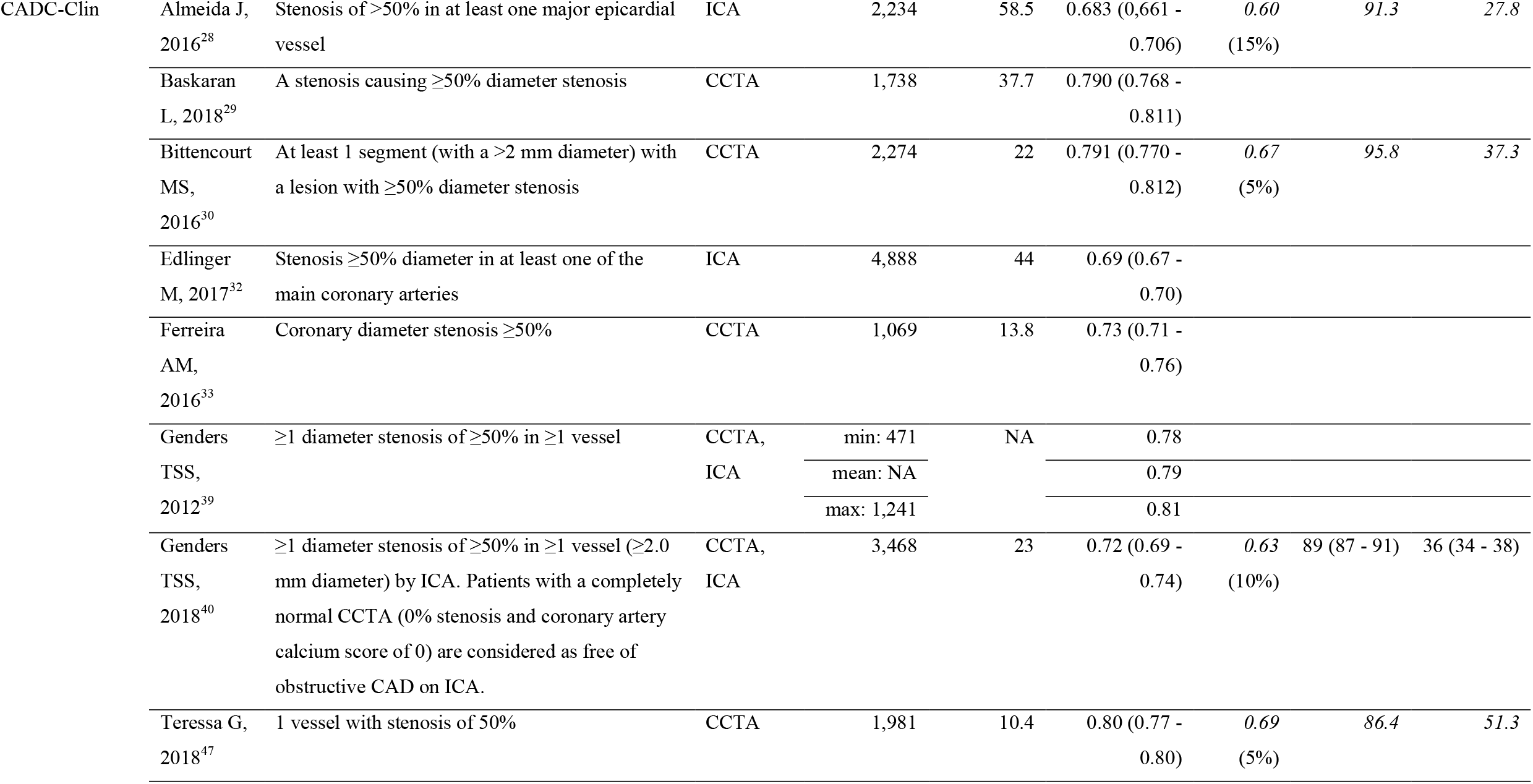
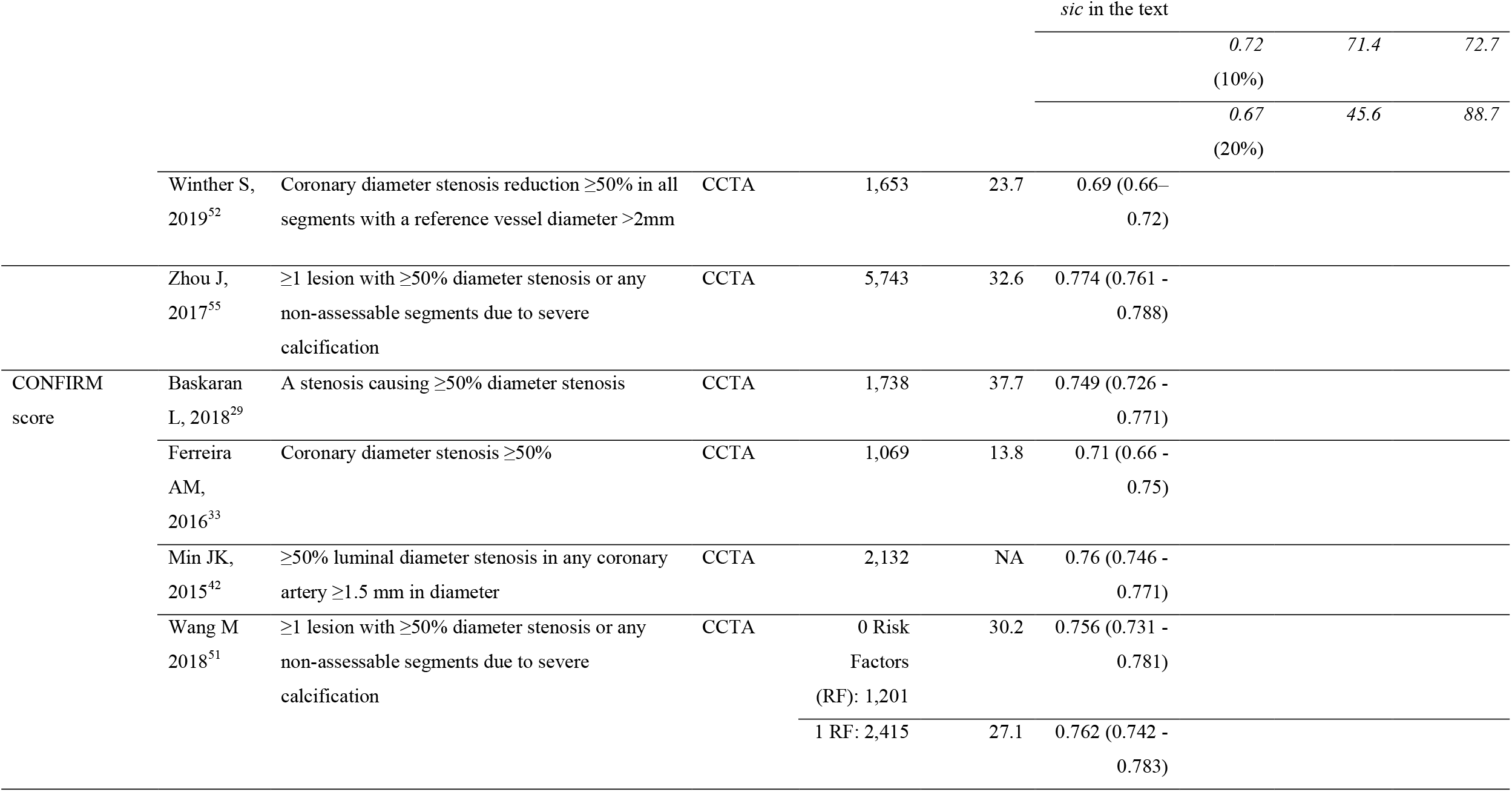
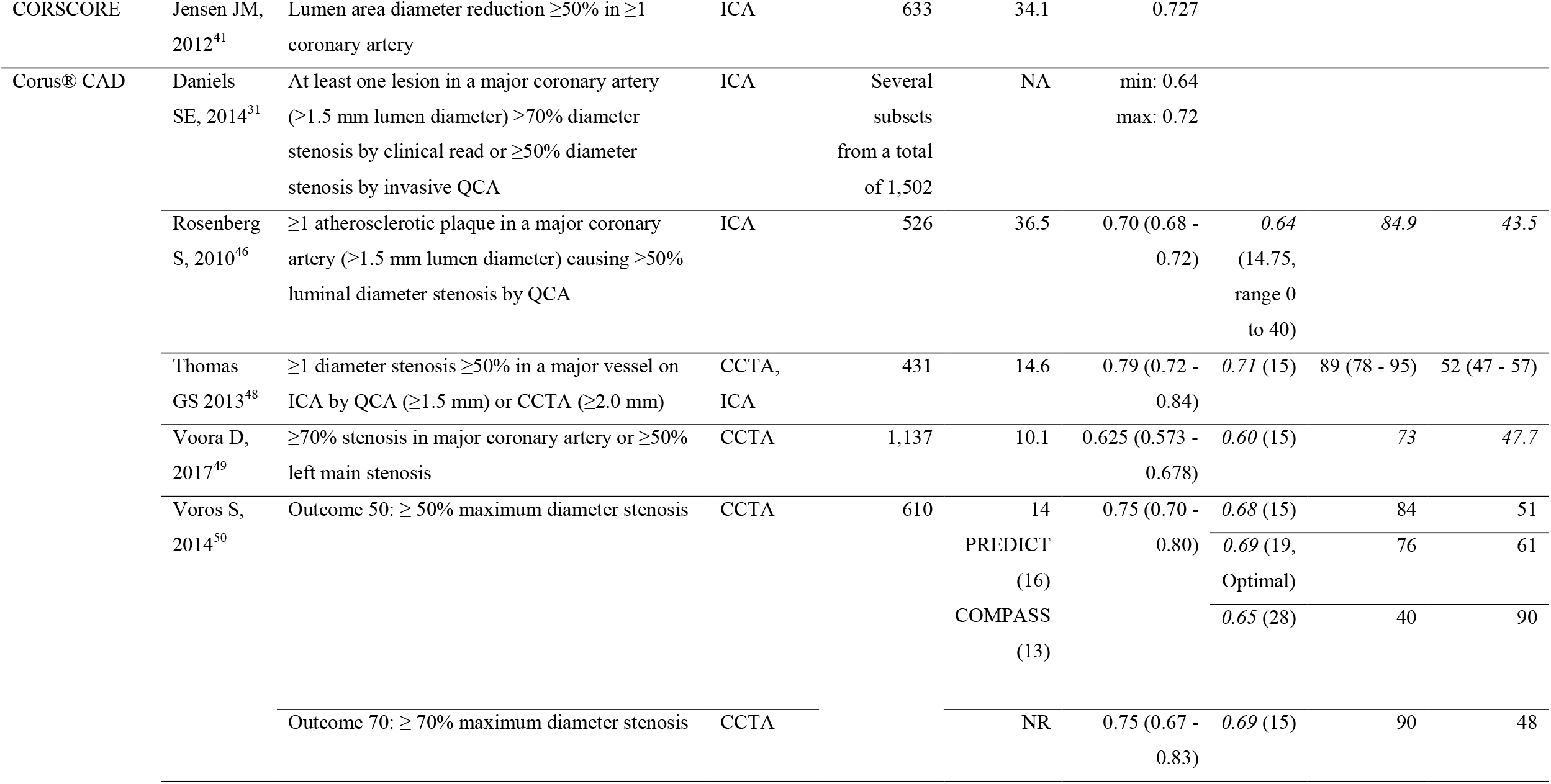
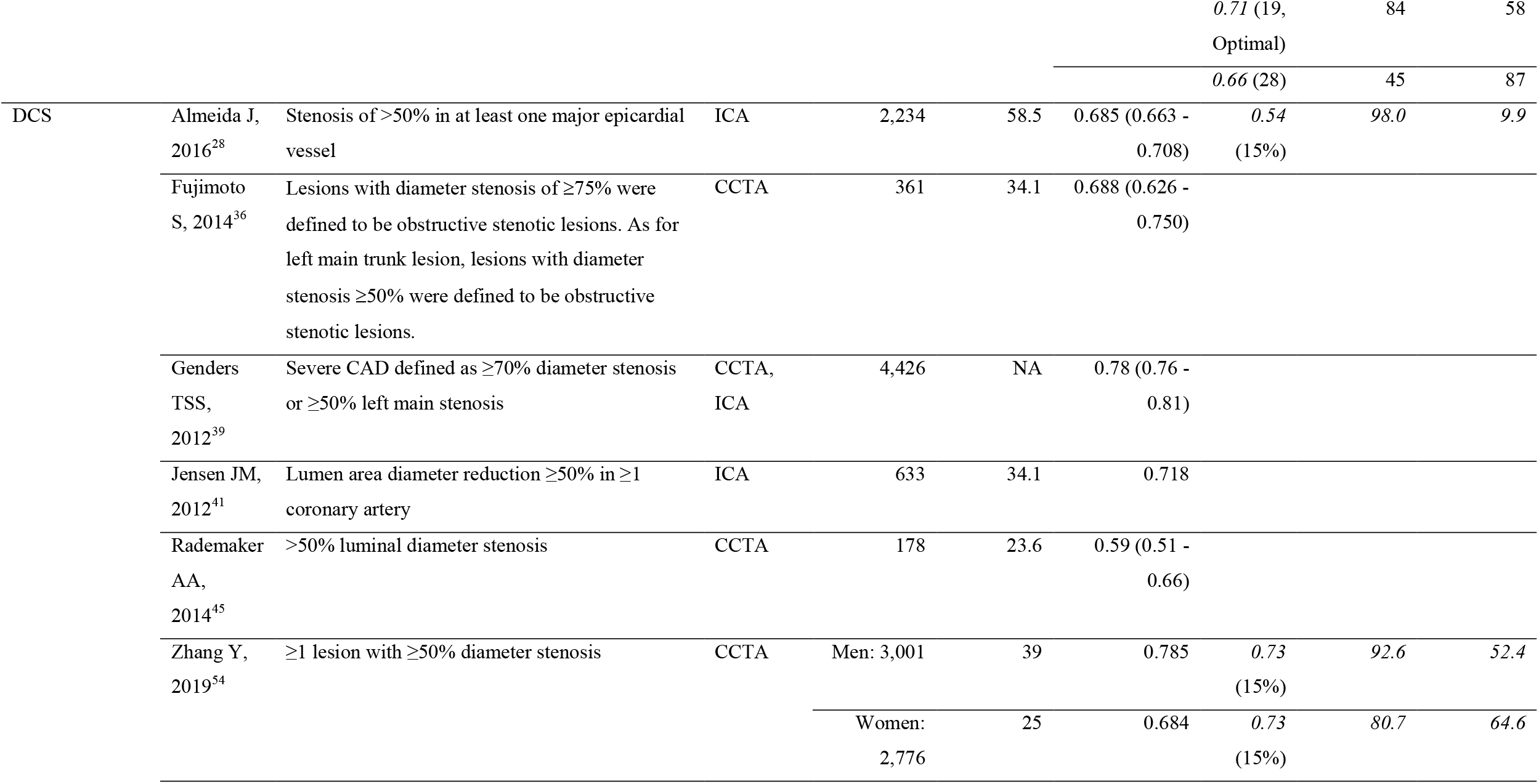
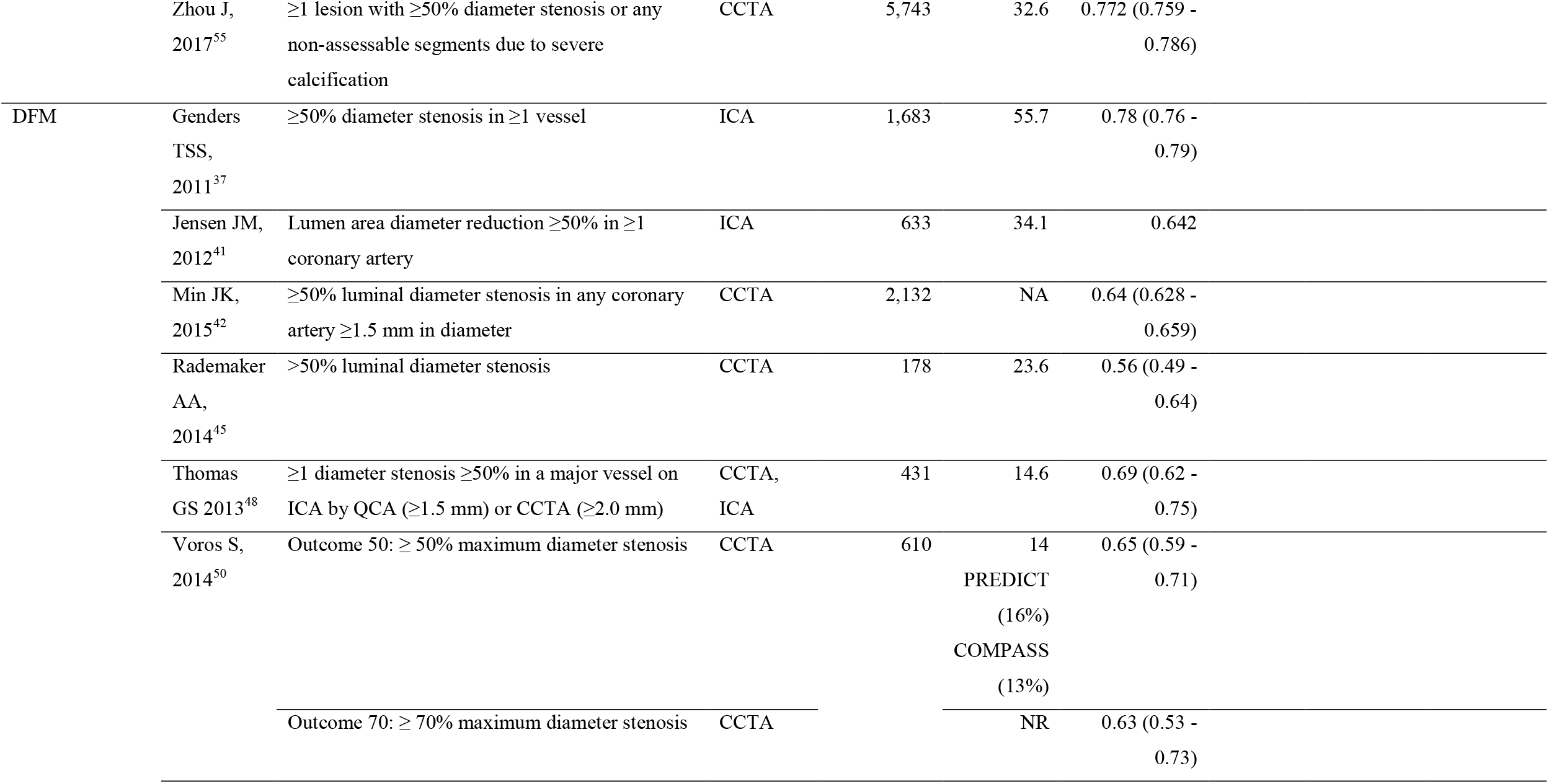
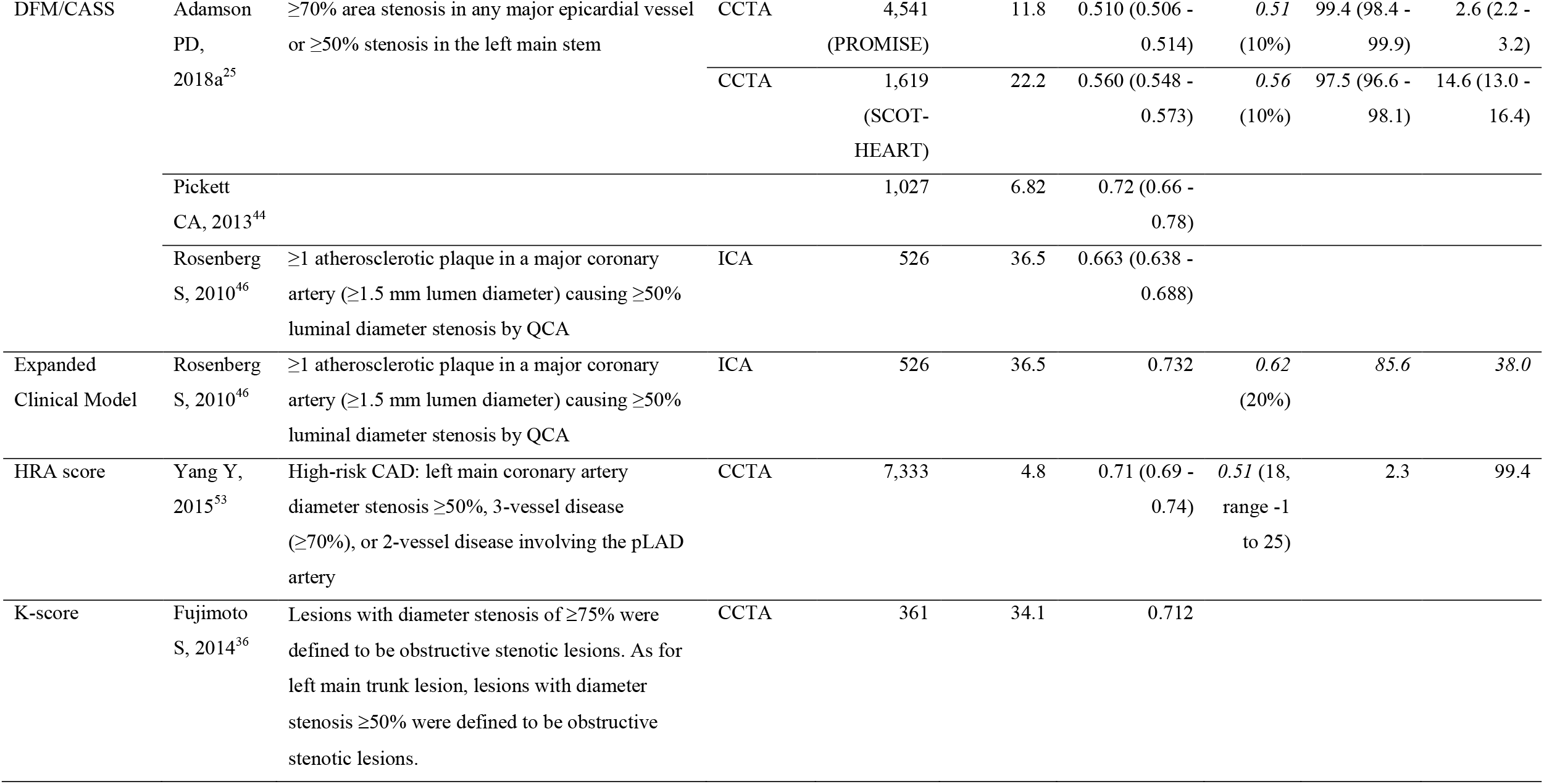
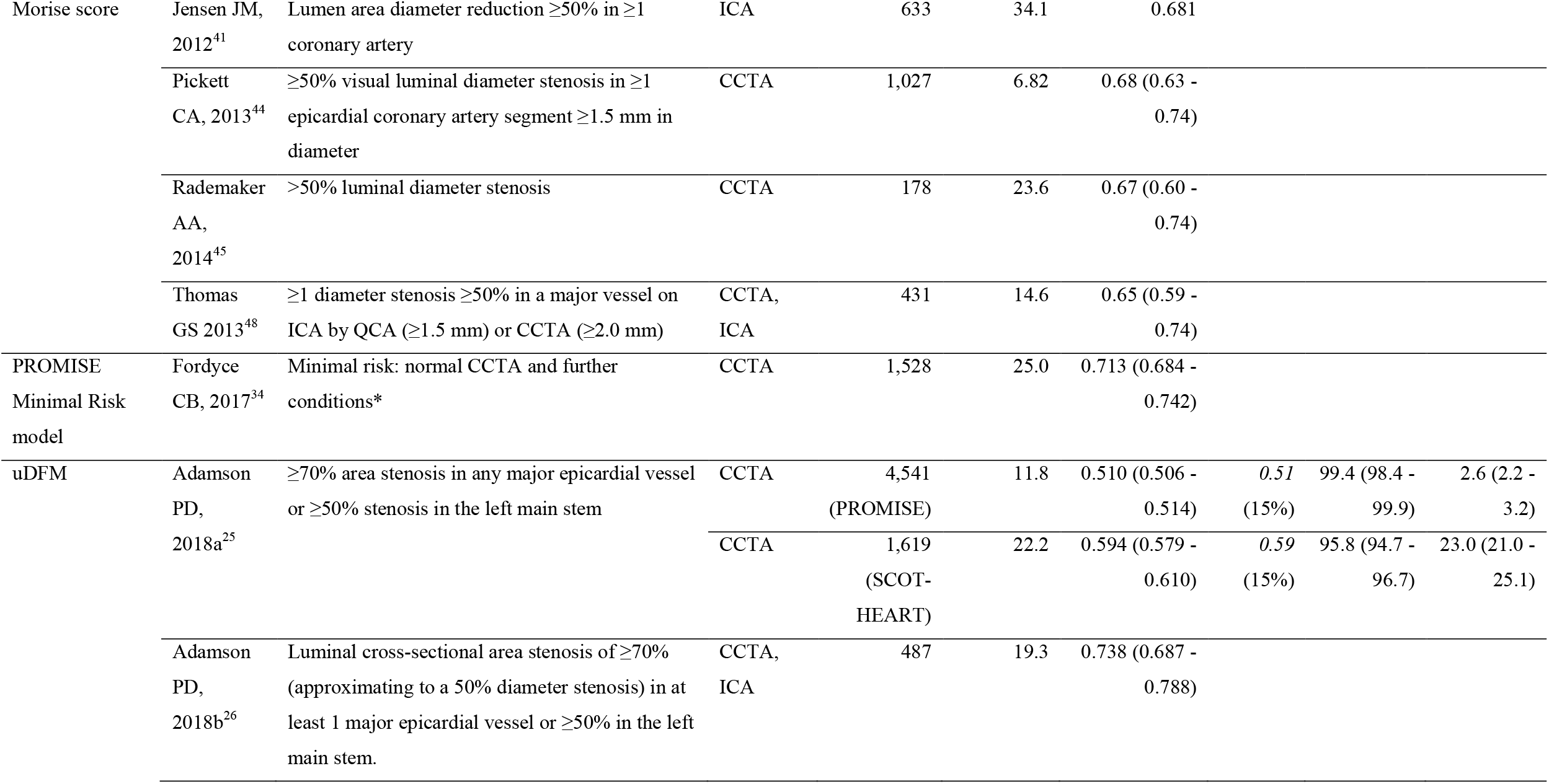
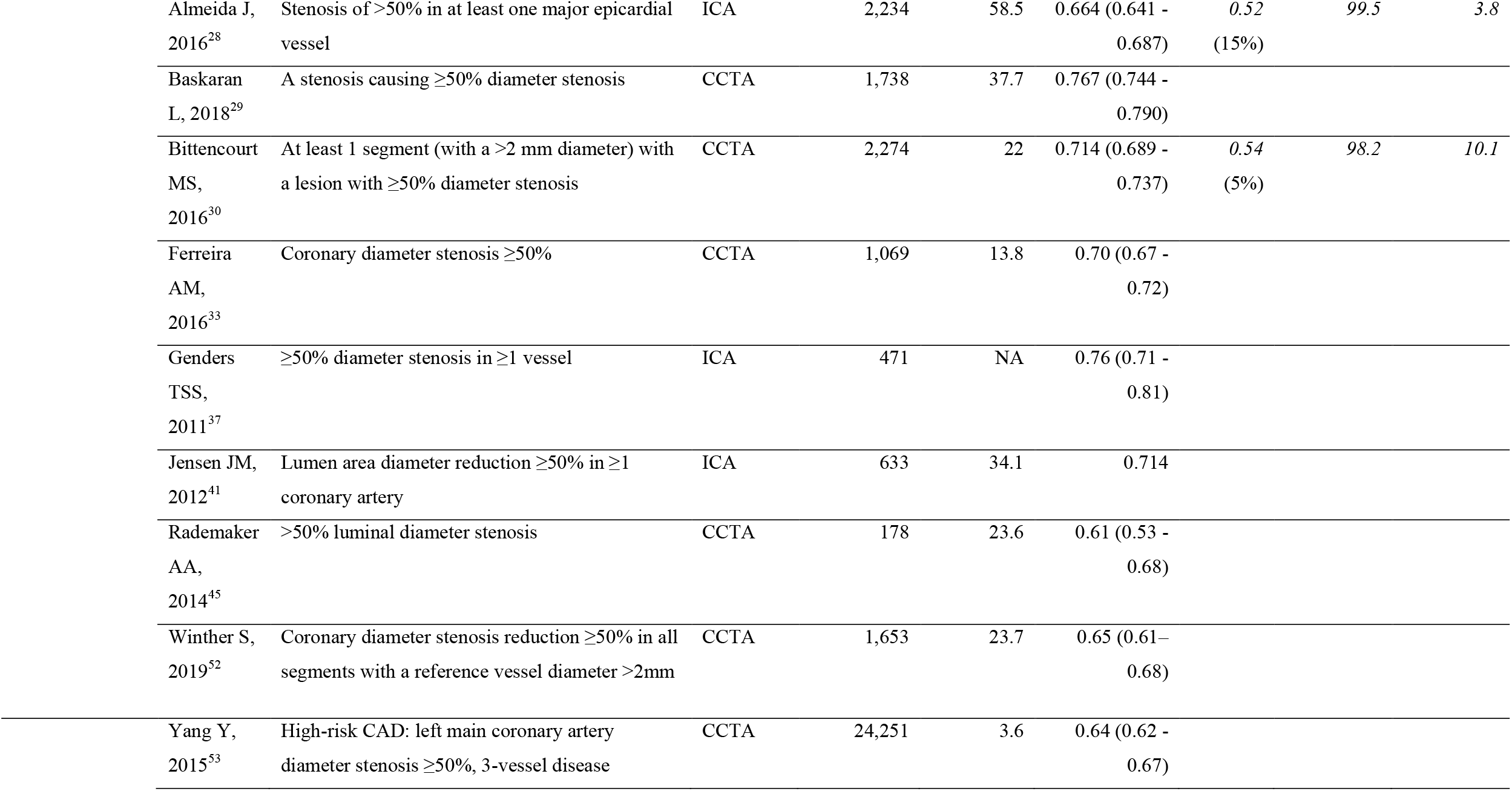
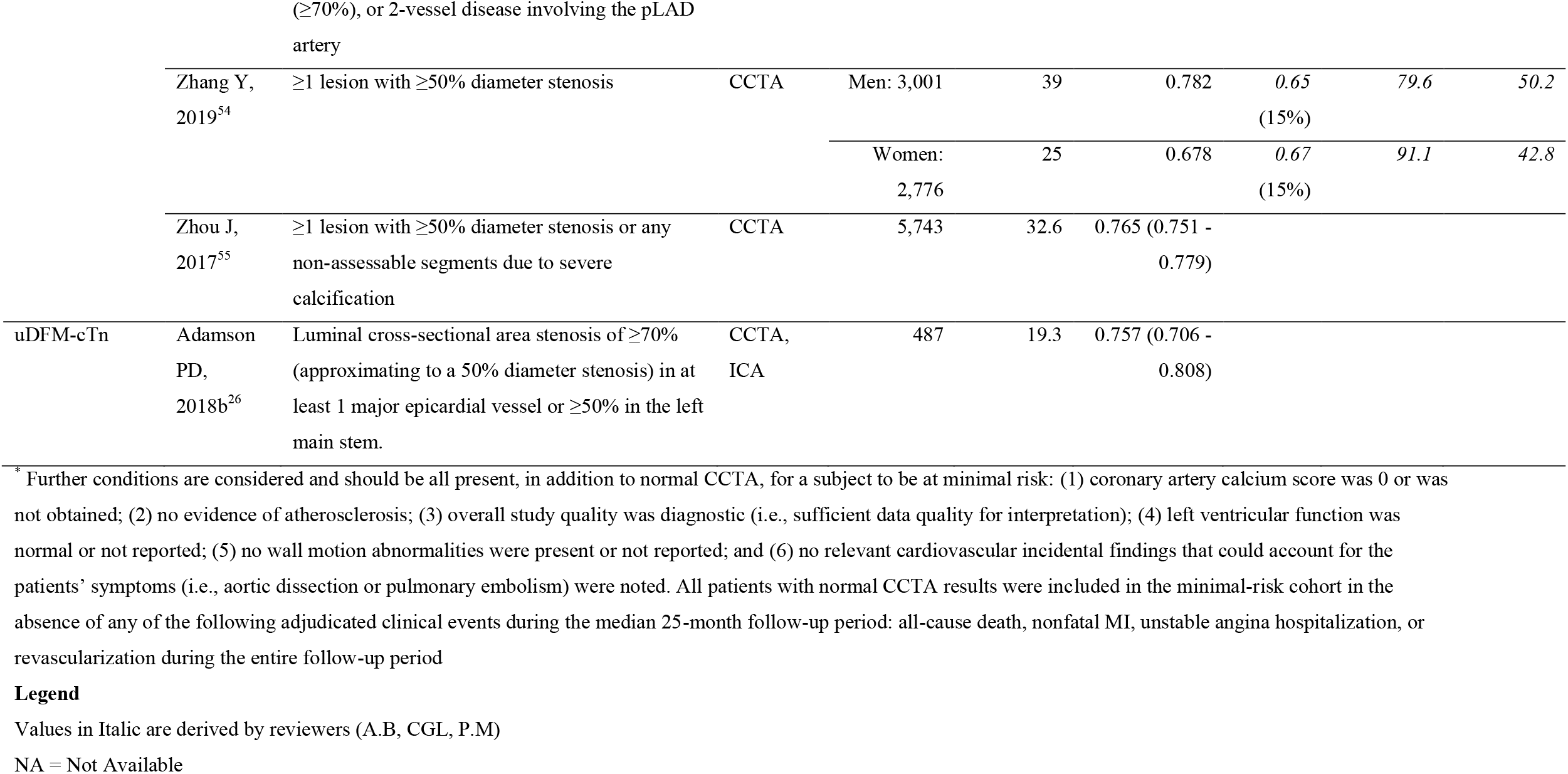
PTP Models performance as reported in the selected studies.

It is worth noting the presence of relevant heterogeneity sources: the high variability of sample sizes (from 178^45^ to 24,251^53^); whether the AUC is computed with respect to a specific PTP cut-off; the adoption of different endpoints. On the one hand, Fordyce et al.^34^ focused on patients unlikely to have CAD, clinical events or revascularisation, who were defined as being at “minimal risk”. Minimal risk is characterised by a normal CCTA and the presence of additional positive conditions. On the other hand, Yang et al.^53^ consider subjects with a high-risk CAD, defined as left main coronary artery diameter stenosis ≥ 50%, 3-vessel disease (diameter stenosis ≥ 70%) or 2-vessel disease involving the proximal left anterior descending coronary artery. Endpoint heterogeneity is also one of the reasons for the large variation of prevalence, from 3.6%^53^ to 58.5%.^28^

AUC values range from 0.51^25^ to approximately 0.81^39^. These data indicate a degree of discriminative performance that varies from almost failing to almost excellent. The CAD-Clin model only has an AUC > 0.80, and this performance level is confirmed in other validations (AUC≥0.79 and 95% confidence intervals [CIs] including 0.80).^29,30,47^ The external validations for the American College of Cardiology Foundation (DFM/CASS model) and ESC guidelines (uDFM)^25^ yield the lowest AUC. This result is not unexpected because these values arise from considering the recommended discriminating cut-offs (10% and 15%, respectively) that directly reflect the preference for high sensitivity. These values cannot be compared to other AUC values that are not derived from fixed cut-off: for ease of comparison, in Table 3 the distinction is made between running and fixed cut-off. All the other models except the Morise score (AUC from 0.65^42^ to 0.68^32,50^) reached a moderate discriminative ability (AUC from 0.70-0.80) when considering the running cut-off AUCs.

The uDFM has been validated on a very different sized populations (from 173 to more than 20,000 subjects) with variable prevalence (from 3.6-58.6%). The most complete validation of the model, considering calibration-in-the-large, recalibration and eventually re-estimation, has been performed by the developers themselves^37^ who obtained a valid overall effect of predictors. The other validating procedures limit themselves to AUC computation and to a rough assessment of under/overestimation, mainly by the Hosmer-Lemeshow goodness-of-fit (HL) test and related calibration plots (calibration-in-the-large is applied in one study^33^). The extension of uDFM with the use of high-sensitivity cardiac troponin I (uDFM-cTn), has a significantly higher AUC than uDFM alone (0.757 versus 0.738, *p*= 0.025) and better calibration HL *p*= 0.0001 versus HL *p*= 0.1123).^25^ The uDFM updated and extended the traditional DFM to a contemporary cohort that included subjects 70 years and older. The CAD Consortium Basic model (CADC-basic) can be considered as a further update on a different contemporary population (see Table 2). With regards to the DFM (and its DFM/CASS version), overestimation is usually reported, especially in women.^45^ Apart from one study,^37^ the DFM was not extensively validated but only used as a usual reference model^41,44,45^ or as a basis to establish the performances of the Corus® CAD model.^46,48,50^

Among the models that include clinical risk factors, DCS and CADC-Clin have been extensively validated. The former generally overestimates prevalence and shows a lack-of-fit by the HL test. Moreover, miscalibration results from a reduced effect of sex and chest pain typicality and an increased effect of diabetes and dyslipidaemia.^39^ The latter has been verified by external validation^32,40,47,52^ Results on miscalibration analysis could be considered quite consistent across papers. This finding indicates smaller than expected effects of the diagnostic characteristics, chest pain typicality in particular.^32,40,47^ Model calibration can be worse in women compared to men, a situation that also arises from the validation of other models (e.g., DFM^37^). The CADC-Clin performances significantly improve with respect to the related CADC-basic.^30,39,40,47^ Comparisons of either uDFM or CADC-Clin with the PROMISE history-based score do not lead to a clear evaluation of the advantages of one over the other in terms of AUC,^29,33^ while the CONFIRM score proves to be better than the DFM.^42^ The substantially steady results of the CONFIRM score on several data-sets are also confirmed on a validation data-set consisting of subjects at the low extreme of traditional cardiovascular risk factor burden.^51^

The Corus® CAD model stands out from the previous ones because it defines an age- and sex-specific gene expression score. Validation is performed by AUC comparisons, HL test and additivity to DFM and other models. The validation procedures show significant AUC improvement when the score is added to other models (e.g., 0.81 versus 0.65 when added to Morise score, with non-overlapping confidence intervals^48^; 0.721 versus 0.663 when added to DFM, *p*= 0.003^46^; not shown in the table). With respect to the Morise score, the only model that explicitly considers a female-specific factor, namely the oestrogen status, the Corus® CAD has significantly higher AUC (0.79 versus 0.65, *p*< 0.001^48^). Testing the Corus® CAD model on different data sets from an extension of the original validation population provides results very similar to the original ones.^31^

Finally, the Minimal Risk model upsets the usual point of view because it aims to directly identify patients with chest pain and normal coronary arteries. Unfortunately, the only other external validation published up to the date of our search^57^ cannot be considered here because it was based on a former version of Fordyce et al. 2017^34^ that includes some computational errors^35^.

With the exception of a few papers that discussed the classical DFM and DCS,^25,28,54^ an in-depth study of the model performances with respect to operational cut-offs is mainly related to the CAD Consortium models and the Corus® CAD model. As far as the CAD Consortium models are concerned, clinical usefulness is assessed at cut-offs that vary from 5%-20%. A cut-off of 14.75 (15 in subsequent works) was identified for the Corus® CAD model in the main work,^46^ a value that corresponds to a disease likelihood of 20% on a validation data set (positivity for index ≤ 15). Overall, sensitivity and specificity values are similar to those we derived for CADC-Clin: both these models show a higher balance between sensitivity and specificity than the guidelines and the DCS on the validation data sets. Finally, as suggested by the high values of sensitivity we derived in Table 3, the low AUC value of the uDFM obtained in Adamson^25^ at the cut-off of 15% has been confirmed by Almeida,^28^ Bittencourt^30^ and, to a lesser extent, Zhang^54^; the corresponding AUC values are 0.52, 0.54 and 0.65 (for men) and 0.67 (for women). Analogous results come from the DCS’s validations.^28,54^

## 4 Validated PTP models: Strengths and weaknesses

External validation is an indispensable tool for investigating the generalisability of a PTP model to populations that differ from the development population study. This process can utilise different approaches, from the computation of indexes to more complex procedures that aim at understanding how the original model should adapt to the new population. The papers included in this review mainly rely on AUC which only allows for a limited comparison among models. Different endpoint definitions and decision on whether or not to adopt a specific cut-off can yield different AUC values, as already highlighted. Moreover, only the whole receiver operating characteristic (ROC) curve will allow evaluation of the clinical usefulness of a test by showing the true positive and false positive fractions that will be obtained for any eventually chosen cut-off. As reported in our results, almost all the models provide a moderate discriminative ability (AUC from 0.70-0.80). Unfortunately, once the model is transferred into an operating scenario and the selection of a specific discriminatory cut-off is required, a clinical protection approach leads clinicians to prefer a very high sensitivity, which of course implies low specificity.^58,59^ Only Corus® CAD,^48,50^ CADC-Clin^47^ and DCS^54^ reach a moderate discriminative level at specified thresholds (0.71, 0.72 and 0.73, respectively). Notably, Corus® CAD recently lost Medicare coverage in the US.^60^

Despite the fact that all the models are obtained by regression techniques, which allow the interpretation of the effect of the predictor on the outcome of interest, very few papers^32,37,40,47^ address a complete validation procedure without rejecting a model after obtaining a poor preliminary performance on the new population by some test. Rather, a different model is developed, without any further in-depth analysis of the failure reason. Regardless of the quality of the new developed model, the lack of adequate consideration of in-depth validation procedures involves the loss of the information captured by the initial study and hinders a deep understanding of how effect size of relevant risk factors can change in a different geographical or setting framework.^24^ For instance, deep validation procedures like miscalibration analysis allow questioning the effect of chest pain typicality in different data sets.^32,40,47^ This finding is consistent with what was recently noted by Di Carli and Gupta:^61^ angina remains a common presenting symptom in a high proportion of cardiac patients that do not show obstructive lesions in their coronary angiograms.

A central question is what clinical cardiologists are most interested in evaluating: CAD of any degree, high-risk plaques, stenosis of a certain anatomic/physiologic severity, stenosis that leads to ischaemia, stenosis that requires intervention or stenosis that must be fixed to reduce adverse outcomes. The answer determines which diagnostic pathway and test is the most appropriate^61,62^ and also affects statistical analysis. A carefully defined outcome should be required to provide a reliable basis for the evaluation of the effect of any predictive variable.^63^ When referring to validation specifically, the application of a model to predict an outcome different from the originally intended one raises some concerns and, eventually, should be explicitly noted. In data-driven models, the outcome definition in the population study also influences predictor selection. Thus, a small AUC value in the validation set does not necessarily indicate a lower performance of the original model on the new population. Instead, it suggests that the model may not be appropriate for the context.^57^

## 5. Conclusions

Several agencies and scientific organizations emphasise the need for increasing the knowledge on how the prediction of the disease can be modified according to the risk factors present in any specific study population or, possibly, in any particular patient. This would indeed improve the precision of the estimated clinical likelihood of CAD. However, the increasing availability of large data-sets, and the highly improved computational power seem to have directed large part of recent researches towards model development rather than model validation.^17^ First of all, our review makes an important selection among the many developed models by mainly considering those externally validated. Then, it provides insights into the effects of traditional and emerging risk factors, biomarkers, and comorbidities on the PTP of obstructive CAD. Finally, our findings lead to the following important recommendations. To achieve a more robust exploitation of PTP models in decision-making processes, significant endpoints should be more clearly stated and consistently measured both in the derivation and validation phases. Furthermore, more comprehensive validation analyses should be adopted to understand model weaknesses. Finally, increased efforts are still needed to thresholds validation and to analyse the effect of PTP on clinical management.

## Data Availability

All data generated or analysed during this study are included in this article and in its supplementary information files.

## Author contributions

AB, CGL, PM, GP and SS provided substantial contribution to the conception of the work. CGL and PM performed the literature search and retrieved selected publications.

All the authors contributed to the extraction and analysis of data. AB, CGL, PM and MRT assessed the quality of included studies.

All the authors contributed to draft the work. AB, CGL and PM revised it critically.

All the authors approved the version to be published and are accountable for all aspects of the work.

CGL is responsible for the overall content as guarantor.

## Acknowledgments

Authors express their great appreciation to Dr. Philip D. Adamson for the valuable discussion on some methodological aspects.

The authors would also like to thank Dr. Tommaso Leo for the clarifications provided on some clinical aspects.

Finally, the authors thank Roberto Guarino (National Research Council of Italy, Institute of Clinical Physiology, Lecce) for his technical support (Informatics Tools for document management and title and abstract screening).

## Supplementary Material

**Additional File 1 - Search strategy** – It is the full search string adopted in OVID. **Additional File 2 – Study design and Eligibility Criteria of main studies** – It provides details on the main studies cited in Table 1.

**Additional File 3 – Proportion of studies with low, high or unclear risk of bias** – It is a summary of the quality assessment according to QUADAS2.

**Additional File 4 – PRISMA Checklist**

