## Additional File 1 for "Validated models for pre-test probability of stable coronary artery disease: a systematic review suggesting how to improve validation procedures"

### Additional FILE 1 - Search strategy

**▼ Search History** (28)

| # ▲ | Searches | Results |
|---|---|---|
| 1 | ▶ Angina Pectoris/ or Angina Pectoris.af. or Angina, Stable/ or (Stable Angina* or Chronic Angina*).af. | 98847 |
| 2 | ▶ Chest Pain/ or Chest Pain*.af. | 72609 |
| 3 | ▶ Coronary Heart Disease/ or (CHD or Coronary Disease* or Coronary Heart Disease*).af. | 412639 |
| 4 | ▶ Coronary Artery Disease/ or (CAD or Coronary Artery Disease* or Coronary Arteriosclerosis or Coronary Atheroscleros*).af. | 288034 |
| 5 | ▶ Coronary Stenosis/ or (Coronary Stenos* or Artery Stenos*).af. | 64518 |
| 6 | ▶ or/1-5 | 770062 |
| 7 | ▶ stratification score*.af. | 492 |
| 8 | ▶ Likelihood Functions/ or Likelihood Function*.af. or (likelihood adj5 disease).af. or CAD likelihood.af. or "predict* CAD".af. | 46652 |
| 9 | ▶ forecasting/ or (pre-test probabilit* or PTP or predictive model* or prediction or forecast*).af. | 664543 |
| 10 | ▶ (probability adj5 disease).af. | 8725 |
| 11 | ▶ or/7-10 | 716518 |
| 12 | ▶ Coronary Angiography/ or Angiograph*.af. | 574196 |
| 13 | ▶ Angiocardiography/ or Angiocardiograph*.af. | 21132 |
| 14 | ▶ Cardiac Catheterization/ or (Cardiac Catheterization* or Heart Catheterization*).af. | 122622 |
| 15 | ▶ Computed Tomography Angiography/ or (Coronary Computed Tomography Angiograph* or CCTA).af. | 21751 |
| 16 | ▶ or/12-15 | 677008 |
| 17 | ▶ 6 and 11 and 16 | 6069 |
| 18 | ▶ (ANIMALS not HUMANS).sh. | 7244023 |
| 19 | ▶ 17 not 18 | 5985 |
| 20 | ▶ limit 19 to english language or limit 19 to italian | 5708 |
| 21 | ▶ remove duplicates from 20 | 2682 |
