## Additional File 2 for "Validated models for pre-test probability of stable coronary artery disease: a systematic review suggesting how to improve validation procedures"

**Additional File 2 – Study design and Eligibility Criteria of main studies**

| | |
|---|---|
| Acronym | **COMPASS** |
| Name | Coronary Obstruction Detection by Molecular Personalized Gene Expression (Corus CAD or ASGES) |
| ClinicalTrials.gov Identifier | NCT01117506 |
| Study design | Observational prospective study. The study enrolled a patient population that presented with stable chest pain syndrome or anginal equivalent and referred for stress myocardial profusion imaging |
| Study Center(s) | Multicenter trial: US (7 centers) |
| Inclusion Criteria: | • Ages 45-90 for women; 35-90 for men.<br>• Stable chest pain syndrome (typical or atypical) or anginal equivalent in the judgment of the investigator (e.g., pain in the neck, jaw, arm or shoulder or dyspnea possibly due to cardiac ischemia).<br>• Referred for a stress test using Myocardial Perfusion Imaging.<br>• The patient has signed the appropriate Institutional Review Board approved Informed Consent Form |
| Exclusion Criteria | • History of known MI or significant CAD.<br>• Current MI or acute coronary syndrome.<br>• Current New York Heart Association (NYHA) class III or IV congestive heart failure symptoms.<br>• Severe regurgitant or stenotic cardiac valvular lesion.<br>• Severe left ventricular systolic dysfunction (LVEF ≤ 35 % documented in the last year); if no assessment was performed or documented in the |

year preceding enrolment, presume normal LVEF.

- Active systemic infection (diagnosed by a combination of clinical symptoms and laboratory testing, including but not limited to fever, leukocytosis, positive blood cultures, pneumonia, urinary tract infection, or abscess in the preceding 2 months) or chronic infection (e.g., HIV, Hepatitis B or C, Tuberculosis).
- Protocol-specified rheumatologic, autoimmune or hematologic conditions (e.g., rheumatoid arthritis, systemic lupus erythematosis, polymyalgia rheumatica, or systemic sarcoidosis).
- Known or suspected diabetes mellitus or documented Hemoglobin A1c (HbA1c) ≥ 6.5; presume normal HbA1c if none documented.
- Total WBC ≥ 11,000 cells/ul and platelet count ≤ 75,000 cells/ul from a CBC with differential drawn within 7 days prior to enrollment [WBC ≥ 11,000 cells/ul and platelet count ≤ 75,000 cells/ul from a CBC drawn > 7 days prior need to be re-drawn at enrollment].
- Recipient of any organ transplant.
- Immunosuppressive or immunomodulatory therapy including any dose of systemic corticosteroids in the preceding 2 months.
- Chemotherapy in the preceding year.
- Major surgery in the preceding 2 months.
- Blood or blood product transfusion in the preceding 2 months.
- Subjects for whom all forms (stress or pharmacologic) of MPI are contraindicated.
- Subjects for whom invasive coronary angiography or coronary CT angiography is contraindicated, including IV beta-blocker.

| | |
|---|---|
| | • Subjects who planned to decline research CCTA or invasive coronary angiography, regardless of MPI result.<br>• Subjects with history of atrial fibrillation/flutter or frequent irregular or rapid heart rhythms.<br>• Known history of renal insufficiency (serum creatinine ≥ 2.0 mg/dL), or severe allergy to iodinated contrast. |

| | |
|---|---|
| Acronym | **CONFIRM** |
| Name | COroNary CT Angiography Evaluation For Clinical Outcomes: An InteRnational Multicenter Registry |
| ClinicalTrials.gov Identifier | NCT01443637 |
| Study design | Observational prospective study. Patients included in the CONFIRM Registry are those that have previously undergone clinically-indicated CCTA as part of their standard of care. |
| Study Center(s) | Multicenter trial: North America, Europe and Asia |
| Inclusion Criteria: | • Age > 18 years<br>• Evaluation by CCTA with 64-detector rows or greater for CAD evaluation as part of standard of care<br>• Interpretable CCTA<br>• Prospective data collection for CAD risk factors. |
| Exclusion Criteria | • No explicit patient exclusion criteria are defined. |

| | |
|---|---|
| Acronym | **PREDICT** |
| Name | Personalized Risk Evaluation and Diagnosis (Using Corus CAD or ASGES) in the Coronary Tree |
| ClinicalTrials.gov Identifier | NCT00500617 |
| Study design | Observational prospective study. The study enrolled patients undergoing clinically indicated invasive coronary artery angiogram or CT angiogram |
| Study Center(s) | Multicenter trial: US |
| Inclusion Criteria: | • Age 21 to 99 Years<br>• Referral for a coronary angiogram (either invasive X-ray angiography or coronary CTA)<br>• Any one of the following clinical syndromes:<br> o chest pain syndrome, stable angina, or anginal equivalent suggesting myocardial ischemia<br> o low-risk unstable angin, or<br> o asymptomatic individuals with a high probability of CAD. |
| Exclusion Criteria | • History of myocardial infarction or known CAD<br>• Current Myocardial Infarct (MI), acute coronary syndrome with high-risk features or unstable angina with high-risk features<br>• New York Heart Association (NYHA) class III or IV congestive<br>• Inability to give informed congestive heart failures<br>• Severe left ventricular systolic dysfunction (LVEF<35%)<br>• Severe regurgitant or stenotic cardiac valve lesion<br>• Active or chronic systemic infection<br>• Rheumatologic, autoimmune or hematologic conditions |

| | |
|---|---|
| | • Any organ transplant<br>• Immunosuppressive therapy<br>• Chemotherapy in the preceding year<br>• Major blood or blood product transfusion in the preceding 2 months |

| | |
|---|---|
| Acronym | **PROMISE** |
| Name | PROspective Multicenter Imaging Study for Evaluation of Chest Pain |
| ClinicalTrials.gov Identifier | NCT01174550 |
| Study design | Interventional (Clinical Trial) randomized study |
| Study Center(s) | Multicenter trial: North America |
| Inclusion Criteria: | • New or worsening chest pain suspicious for clinically significant coronary artery disease (CAD)<br>• no prior evaluation for this episode of symptoms<br>• planned non-invasive testing for diagnosis<br>• men age ≥55 years<br>• men age ≥45 years with increased probability of coronary artery disease (CAD) due to either (A. Diabetes Mellitus (DM) requiring medical treatment OR Peripheral Arterial Disease (PAD) defined as documented >50% peripheral arterial stenosis treated medically or invasively OR cerebrovascular disease (stroke, documented > 50% carotid stenosis treated medically or invasively) OR B. At least one of the following cardiovascular risk factors: 1-Ongoing tobacco use, 2-Hypertension, 3- |

| | |
|---|---|
| | Abnormal ankle brachial index (ABI) defined as less than <0.9, 4-Dyslipidemia<br>• women age ≥65 years<br>• women age ≥50 years with increased probability of coronary artery disease (CAD) due to either (A. Diabetes Mellitus (DM) requiring medical treatment OR Peripheral Arterial Disease (PAD) defined as documented >50% peripheral arterial stenosis treated medically or invasively OR cerebrovascular disease (stroke, documented > 50% carotid stenosis treated medically or invasively) OR B. At least one of the following cardiovascular risk factors: 1-Ongoing tobacco use, 2-Hypertension, 3-Abnormal ankle brachial index (ABI) defined as less than <0.9, 4-Dyslipidemia<br>• Serum creatinine ≤ 1.5 mg/dL within the past 90 days<br>• Negative urine/serum pregnancy test for female subjects of child-bearing potential |
| Exclusion Criteria | • Diagnosed or suspected acute coronary syndrome (ACS) requiring hospitalization or urgent or emergent testing; Elevated troponin or creatine kinase-myocardial band (CK-MB)<br>• Hemodynamically or clinically unstable condition systolic blood pressure (BP) < 90 mmHg, atrial or ventricular arrhythmias, or persistent resting chest pain felt to be ischemic despite adequate therapy)<br>• Known coronary artery disease (CAD) with prior Myocardial infarction (MI), percutaneous coronary intervention (PCI), coronary artery bypass graft (CABG) or any angiographic evidence of coronary artery disease |

| | |
|---|---|
| | (CAD) ≥50% lesion in a major epicardial vessel<br>• Any invasive coronary angiography or non-invasive anatomic or functional cardiovascular (CV) test for detection of coronary artery disease (CAD), including coronary tomographic angiography (CTA) and exercise electrocardiogram (ECG), within the previous twelve (12) months<br>• Known significant congenital, valvular (> moderate) or cardiomyopathic process (hypertrophic cardiomyopathy or reduced systolic left ventricular function (LVEF) ≤ 40%)) which could explain cardiac symptoms<br>• Contraindication to undergoing a coronary tomographic angiography (CTA), including but not limited to: a. Allergy to iodinated contrast agent, b. Unable to receive beta blockers unless heart rate < 65 beats per minute, c. Pregnancy<br>• Life expectancy < 2 years<br>• Unable to provide written informed consent or participate in long-term follow-up |

| | |
|---|---|
| Acronym | **SCOT-HEART** |
| Name | Scottish COmputed Tomography of the HEART Trial |
| ClinicalTrials.gov Identifier | NCT01149590 |
| Study design | Interventional (Clinical Trial) randomized study |
| Study Center(s) | Multicenter trial: Scotland (UK) |

| | |
|---|---|
| Inclusion Criteria: | • 18 and ≤75 years of age<br>• Attendance at the Rapid Access Chest Pain Clinic |
| Exclusion Criteria | • Inability or unwilling to undergo computed tomography scanning, such as exceeding weight tolerance of scanner<br>• Severe renal failure (serum creatinine >200 µmol/L or estimated glomerular filtration rate <30 mL/min)<br>• Previous recruitment to the trial<br>• Major allergy to iodinated contrast agent<br>• Unable to give informed consent<br>• Known pregnancy<br>• Acute coronary syndrome within 3 months |
