## Supplementary figures and images for "Validated models for pre-test probability of stable coronary artery disease: a systematic review suggesting how to improve validation procedures"

### Additional File 3

**Additional File 3 – Proportion of studies with low, high or unclear risk of bias**

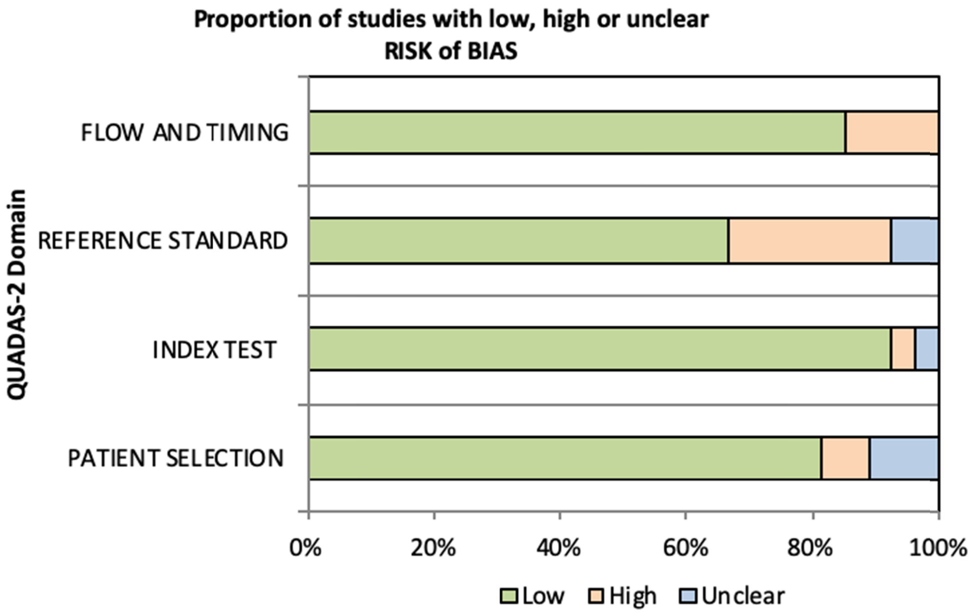
